# Postnatal signals for later cardiovascular morbidity after preterm pre-eclampsia

**DOI:** 10.64898/2026.04.20.26351347

**Authors:** Amy Leslie, Sela Maadh, Megan Lee, Olivia Jones, Lucy Priestner, Kate Duhig, John Farrant, David Hutchings, Josephine Naish, Christopher A Miller, Jenny Myers, Laura Ormesher

## Abstract

**Introduction:** Preterm pre-eclampsia is associated with increased risk of later cardiovascular disease. This study examines cardiometabolic health 3-6 years post-preterm pre-eclampsia and explores whether early postnatal cardiovascular phenotypes relate to later cardiovascular morbidity.

**Methods:** PICk-UP trial participants who experienced preterm pre-eclampsia underwent assessments including anthropometry, blood pressure (BP), arteriography, echocardiography, biomarkers and cardiac magnetic resonance (CMR) imaging 3-6 years postpartum. The primary outcome was hypertension prevalence, with secondary outcomes including cardiac fibrosis, remodelling, and function, obesity, and lipid abnormalities. Associations between baseline, pregnancy and postnatal characteristics with the primary and secondary outcomes were explored.

**Results:** Forty-five women were included; 37 underwent echocardiography and 20 had CMR. At 3-6 years, 53% had hypertension, 32% developed de novo hypertension, 30% had adverse left ventricular (LV) remodelling, 49% had diastolic dysfunction, and 27% were obese. Myocardial fibrosis was detected in 35% of CMR participants. No cardiovascular measures changed from 6 months postpartum to 3-6 years. Women who developed hypertension demonstrated higher BP and LV mass index, from 6 weeks postpartum, with distinct postnatal BP trajectories. Women with myocardial fibrosis exhibited higher sFlt and CRP concentrations from 6 weeks postpartum, with sFlt correlating with native T1 at 3-6 years.

**Discussion:** Women with prior preterm pre-eclampsia show significant cardiometabolic morbidity 3-6 years postpartum. Early postnatal phenotypes indicate long-term cardiovascular risk. Persistent anti-angiogenic imbalance and inflammation may contribute to myocardial fibrosis. Early BP, weight, and biomarker measurement may help identify at-risk women, warranting further studies on optimising postnatal care to mitigate cardiovascular risk after preterm pre-eclampsia.

## Background

Cardiovascular disease (CVD) remains the leading cause of mortality among women worldwide, placing a significant burden on healthcare services and the wider economy, with an estimated annual cost of approximately £29 billion in the UK.^1,2^ Despite this, CVD in women is frequently under-recognized, under-diagnosed, and under-treated, contributing to preventable morbidity and mortality.^3^

This issue is particularly concerning for women with a history of pre-eclampsia, who face a markedly elevated long-term cardiovascular risk. ^4–10^ Pre-eclampsia is a hypertensive disorder of pregnancy affecting approximately 3-5% of pregnancies worldwide. It is defined by new or worsening hypertension (≥140mmHg systolic or ≥90mmHg diastolic) from 20 weeks’ gestation, accompanied by proteinuria and/or evidence of maternal organ/placental dysfunction^11^. Increasing evidence from large observational studies have demonstrated that women with a history of pre-eclampsia have approximately a twofold risk of CVD, including heart failure, ischaemic heart disease and stroke,^5,12,13^ with risk evident within the first year postpartum and being particularly pronounced among those with preterm pre-eclampsia (delivery <37 weeks).^5^

Although delivery of the placenta remains the definitive treatment for pre-eclampsia, it does not mitigate the long-term cardiovascular risk that persists after pregnancy.^14^ Although the mechanisms linking pre-eclampsia to CVD are not fully understood, two broad, non-mutually exclusive hypotheses have been proposed.^15,16^ One suggests that pregnancy acts as a physiological cardiovascular stress test, unmasking pre-existing susceptibility to CVD, as indicated by preeclampsia.^17^ While pre-eclampsia shares several risk factors with CVD, including obesity, insulin resistance, and pre-existing hypertension. In epidemiological studies, the association between pre-eclampsia and later CVD persists even after adjustment for these factors, suggesting that shared risk does not fully explain the relationship.^13,18–21^ An alternative hypothesis proposes that pre-eclampsia itself contributes causally to future CVD, through mechanisms such as persistent angiogenic imbalance, systemic inflammation, endothelial and microvascular dysfunction, and adverse cardiac remodelling.^22–24^

Irrespective of the direction of causality, pregnancy and the early postnatal period represent a unique and under-utilized opportunity for cardiovascular risk assessment and prevention. Leveraging the well-established association between pre-eclampsia and subsequent CVD, and targeting the early postnatal period as a window for intervention, offers an opportunity to identify high-risk women earlier and implement preventative strategies before overt disease develops. Such an approach has the potential to reduce long-term cardiovascular morbidity, while offering a more time-and cost-effective strategy than treating established CVD. Moreover, identifying women at highest risk of subsequent CVD will enable more precise risk stratification and provide mechanistic insights to inform targeted preventive and therapeutic strategies.

This study aimed to investigate an important evidence gap by extending follow-up of the cohort of women with preterm pre-eclampsia established in the PICk-UP study.^25^ At 3-6 years postpartum, we compared baseline characteristics, index pregnancy features, and early postnatal cardiovascular phenotypes between women who did and did not subsequently develop adverse cardiovascular outcomes, including hypertension and myocardial fibrosis. We also describe the prevalence of cardiovascular abnormalities at 3-6 years following preterm pre-eclampsia. This exploratory analysis aimed to identify early features associated with later cardiovascular morbidity, thereby refining risk stratification and informing hypothesis generation for adequately powered prospective studies.

## Methods

### Study design

The PONCHOS (POstNatal Cardiovascular Health OutcomeS) study was a single-centre prospective observational cohort study, following up the PICk-UP (Postnatal Enalapril to Improve Cardiovascular function following preterm Preeclampsia) cohort (NCT03466444; 18/NW/0253).^25^ Participants with preterm pre-eclampsia had previously been randomized to 6 months’ postnatal treatment with enalapril or placebo, or had been enrolled in the PICk-UP observational sub-study and therefore not been randomized to a postnatal intervention. Further details about the PICk-UP study can be found in the original manuscripts.^25,26^ All PONCHOS assessments were conducted at the Maternal and Fetal Health Research Centre, St Mary’s Hospital, Manchester, UK, except cardiovascular magnetic resonance (CMR), which was performed at the BHF Manchester Centre for Heart and Lung Magnetic Resonance Research, Wythenshawe Hospital, UK. All study procedures were approved by the UK Health Research Authority and Regional Research Ethics Committee (REC reference 22/WM/0085), and all participants provided written informed consent. For the present analysis, longitudinal data from PICk-UP (3 visits at 0-3 days, 6 weeks and 6 months’ postpartum) and PONCHOS (3-6 years postpartum) were integrated. Figure 1 provides an overview of study design.

**Figure 1.**
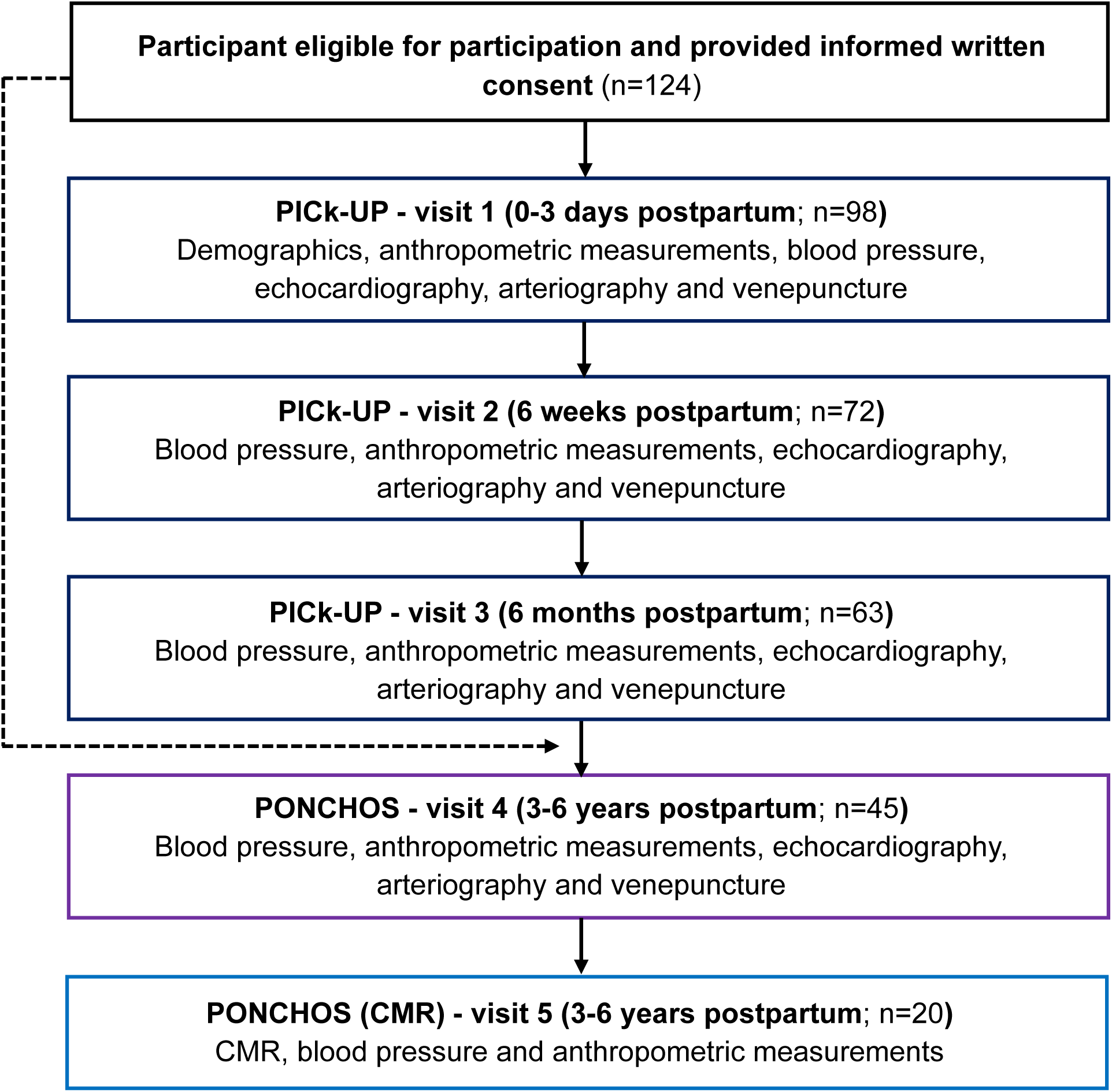
Schematic of the cohort study timeline. Timeline showing recruitment following preterm pre-eclampsia, early postnatal phenotyping, and subsequent longer-term cardiovascular follow-up within linked studies. PICk-UP, Postnatal enalapril to Improve Cardiovascular fUnction following preterm Pre-eclampsia; PONCHOS, POstNatal Cardiovascular Health OutcomeS; CMR, cardiac magnetic resonance imaging.

### Eligibility, recruitment, and cohort derivation

Women aged ≥18 years with an index pregnancy complicated by preterm pre-eclampsia (delivery before 37 weeks’ gestation) who had previously participated in PICk-UP and consented to future contact were eligible for inclusion.^25,26^ Participants were re-contacted by telephone and email and invited to attend a follow-up visit at 3-6 years postpartum. Women were excluded if they were currently pregnant, had known cardiac disease prior to the index pregnancy (as per PICk-UP exclusion criteria), were unable to provide informed consent, or had contraindications to CMR for the nested CMR sub-study.

### Study Procedures

Details of study procedures at the first three postnatal visits are described in the original manuscript.^25^ At the initial 3-6-year PONCHOS visit, women underwent bedside cardiometabolic assessment including: anthropometric measurements (height and weight), BP assessment (in triplicate from the right arm, whilst seated after ≥ 10 minutes’ rest, with the arm supported at the heart level using a validated automated sphygmomanometer: Microlife/Alere, Cheshire, UK), arterial stiffness evaluation using the Tensioclinic Arteriograph (TensioMed, Budapest, Hungary; triplicate measures), transthoracic echocardiography (Voluson E-series, GE Healthcare, to assess left ventricular [LV] remodelling, systolic and diastolic function), and venous blood sampling. A subgroup of eligible participants attended a single additional visit for comprehensive CMR. All measurements were performed according to pre-specified protocols and replicated across visits wherever feasible to ensure valid longitudinal assessment. Further details about study procedures are included in the supplementary materials. Clinical definitions used in this study are detailed in the supplementary material.

### Primary and secondary outcomes

The primary outcome was prevalence of hypertension 3-6 years after preterm pre-eclampsia. The secondary outcomes included the prevalence of: i) de novo hypertension, ii) myocardial fibrosis, iii) cardiac systolic dysfunction, iv) diastolic dysfunction, v) cardiac remodelling, vi) obesity, and vii) lipid abnormalities 3-6 years following preterm pre-eclampsia. Analyses investigated the association between baseline, index pregnancy, and early postnatal characteristics and the primary and secondary outcomes.

### Statistical analysis

Statistical analyses were performed using Stata (StataCorp, College Station, TX, USA). Continuous variables were assessed for distribution and summarized as mean ± standard deviation (SD) or median [interquartile range], as appropriate. Categorical variables were presented as counts (percentages). Prevalence of abnormalities (including hypertension, de novo hypertension, myocardial fibrosis, cardiac systolic dysfunction, diastolic dysfunction, cardiac remodelling, obesity, lipid abnormalities) were reported descriptively.

This study was not powered to formally predict binary cardiovascular outcomes at 3-6 years. Therefore, rather than modelling predictive associations, analyses focused on exploratory comparisons of baseline characteristics, index pregnancy features, and early postnatal cardiovascular measures between women who did and did not subsequently develop cardiovascular morbidity (hypertension and/or myocardial fibrosis) by 3-6 years postpartum.

Group comparisons for continuous variables were performed using the Wilcoxon rank-sum test, and Fisher’s exact test was used to compare categorical variables between groups (defined by 3-6-year hypertension or myocardial fibrosis status). Postnatal cardiovascular phenotypes were additionally compared visually between groups using radar plots. Comparisons between postnatal timepoints were undertaken using the Wilcoxon rank-sum test.

Longitudinal mixed-effects models were used to account for repeated observations within individuals and to explore differences in change over time between groups, defined by cardiovascular morbidity status at 3-6 years. A random effect was specified at the participant level to account for within-individual correlation across repeated measurements. For descriptive purposes, group-level mean values with standard errors were plotted across study visits, either for the full cohort or stratified by 3-6-year cardiovascular hypertension or myocardial fibrosis status.

Associations between continuous measures were assessed using linear regression, with effect estimates reported with confidence intervals. All statistical tests were two-sided, and a p value <0.05 was considered statistically significant.

## Results

### Recruitment

Of the 98 women who completed PICk-UP, 96 consented to future contact. Of these, 45 women consented to participation in PONCHOS and underwent detailed bedside cardiovascular phenotyping at 3-6 years postpartum (Supplementary Figure 1). Thirty-seven women completed echocardiography and 20 underwent CMR imaging. This study was underpowered to explore the long-term impact of short-term postnatal enalapril. Only thirteen women who were randomized to 6 months’ treatment with enalapril completed PICk-UP and attended the 3-6-year PONCHOS visit. Consequently, analyses are presented with all study groups combined (enalapril, placebo and observational).

### Cohort characteristics

Baseline characteristics of the cohort are summarized in Table 1. Supplementary Table 1 demonstrates that the PONCHOS cohort and the CMR subgroup were broadly representative of the original PICk-UP cohort. However, participants in the CMR subgroup were older and less likely to be smokers than the PONCHOS participants who did not have CMR imaging (41.5±4.9 versus 34.4±6.3 years and 5% versus 16% smokers). Forty-four per cent of the cohort were from ethnic minority backgrounds. In this cohort, 53% had fetal growth restriction (FGR; birthweight centile < 3^rd^) and 38% delivered before 34 weeks’ gestation in their index pregnancy.

**Table 1:**
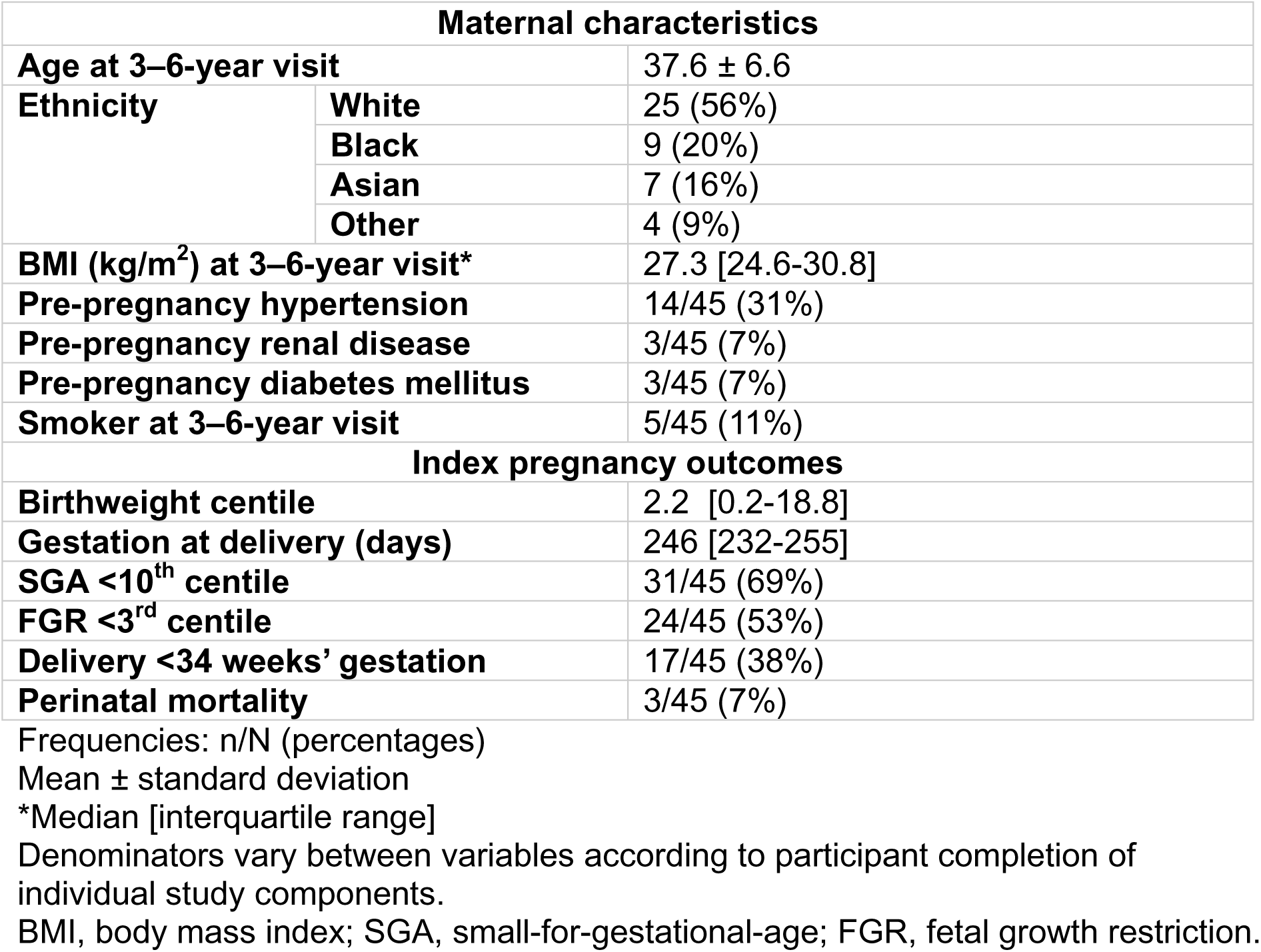
Cohort characteristics (n=45)

### Change in cardiovascular parameters over time

Changes in cardiovascular parameters across timepoints are illustrated in Figure 2, with the greatest magnitude of change occurring within the first 6 weeks postpartum. All cardiovascular measures did not change significantly between 6 months and 3-6 years. BP, weight and indices of diastolic function (E/E’) did not change significantly between 6 weeks and 3-6 years (Supplementary Table 2).

**Figure 2:**
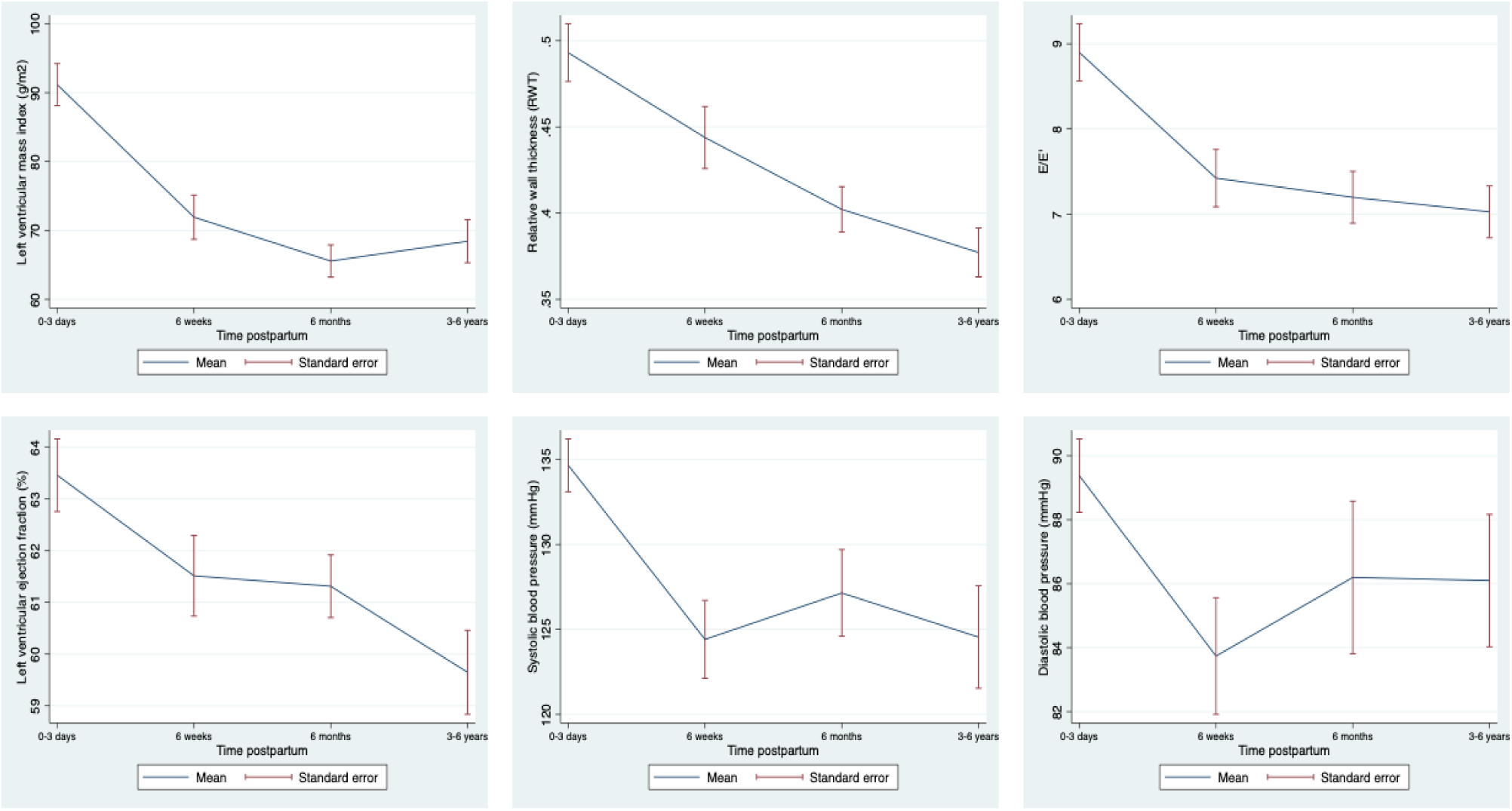
Change in cardiovascular parameters over time. Change in cardiovascular parameters over time: **(A)** left ventricular mass index, **(B)** relative wall thickness, **(C)** diastolic function (E/E′), **(D)** systolic function (left ventricular ejection fraction), **(E)** systolic blood pressure and **(F)** diastolic blood pressure . Lines represent mean values and error bars represent standard errors. Models adjusted for repeated measures within individuals.

### Prevalence of abnormalities at 3-6 years postpartum

A high burden of cardiovascular and metabolic abnormalities was observed 3-6 years following preterm pre-eclampsia (Table 2). Twelve women (27%) were obese, a prevalence unchanged from the start of the index pregnancy. De novo hypertension had developed in 32% of the cohort. Among women who underwent CMR imaging, 35% (7/20) demonstrated evidence of myocardial fibrosis.

**Table 2:** Prevalence of abnormalities at 3-6 years.

|  |  |  |
| --- | --- | --- |
| **Hypertension** |  | 24/45 (53%) |
| **De novo hypertension** |  | 10/31 (32%) |
| **Diastolic dysfunction (BSE)** | **Any** | 18/37 (49%) |
|  | **Stage 1** | 12/37 (32%) |
|  | **Stage 2** | 1/37 (3%) |
|  | **Stage 3** | 5/37 (14%) |
| **Remodelling** | **Any** | 11/37 (30%) |
|  | **Concentric remodelling** | 7/37 (19%) |
|  | **Concentric hypertrophy** | 2/37 (5%) |
|  | **Eccentric hypertrophy** | 2/37 (5%) |
| **Systolic impairment (LVEF < 55%)** | **Any** | 5/37 (14%) |
| **QRISK3 relative risk $\geq 2$** |  | 17/40 (43%) |
| **Fibrosis** |  | 7/20 (35%) |
| **Metabolic** |  |  |
| **Obesity (BMI $\geq 30 \text{ kg/m}^2$)** |  | 12/45 (27%) |
| **Abnormal lipids** |  | 37/45 (82%) |

Frequencies: n/N (percentages).
Denominators vary between variables according to participant completion of individual study components.
BSE, British Society of Echocardiography; QRISK3 relative risk, 10-year cardiovascular risk score relative to age-, sex, and ethnicity-matched individuals with optimal risk factors; LVEF, left ventricular ejection fraction; BMI, body mass index.

Notably, among women who developed myocardial fibrosis, 5/7 (71%) had no significant medical history (including hypertension and diabetes) prior to the index pregnancy. Supplementary Figure 2 illustrates the overlap between abnormalities detected by echocardiography, CMR, BP and biomarker measurements: 5/7 women with fibrosis had at least one additional abnormality detected by bedside cardiovascular assessment (including echocardiography and oscillometric BP measurement). Six out of seven women with fibrosis exhibited raised serum sFlt levels (≥93.30 pg/mL, ranging from 95-104 pg/mL), using Kvehaugen et al.’s reference range derived from women 5-8 years following a normotensive pregnancy.^23^

### Do baseline maternal characteristics relate to cardiovascular health at 3-6 years?

Women who were hypertensive at 3-6 years had higher BP and BMI at booking in the index pregnancy compared with women who remained normotensive (sBP: 127.0 [118.0-138.0] versus 108.0 [100.0-120.0] mmHg, p=0.0002; dBP: 79.0 [70.0-87.0] versus 64.0 [60.0-70.0] mmHg, p<0.0001; BMI: 29.3 [24.6-32.0] vs 23.9 [20.9-27.2] kg/m^2^; p=0.003).

After exclusion of women with pre-existing hypertension, all clinical characteristics at booking were comparable between those who developed de novo hypertension by 3-6 years and those who did not, except diastolic BP (70.0 [70.0-78.0] mmHg versus 64.0 [60.0-70.0] mmHg; Supplementary Table 3). There was a trend towards a higher booking BMI in those who later developed myocardial fibrosis (29.9 [26.9-30.8] kg/m^2^ vs 24.4 [23.9-24.8] kg/m^2^; p=0.05). No other baseline characteristics differed between the fibrosis groups (Supplementary Table 4).

### Do pregnancy outcomes relate to cardiovascular health at 3-6 years?

There was no difference in pregnancy phenotype (including gestation at delivery, birthweight centile, prevalence of FGR, SGA, or preterm birth <34 weeks’) between those who later developed hypertension and those who did not (Supplementary Table 5). However more women who had myocardial fibrosis at 3-6 years had a preterm birth before 34 weeks’ gestation, than those who did not develop fibrosis (71% versus 23%, p=0.04)

### How do early postnatal parameters relate to future cardiovascular morbidity?

Women who were hypertensive at 3-6 years postpartum (24/45) demonstrated distinct cardiovascular phenotypes in the early postnatal period (Figure 3). Compared to women without hypertension, women with hypertension at 3-6 years had higher BP and altered LV remodelling (higher left ventricular mass index [LVMi] and relative wall thickness [RWT]) from 6 weeks postpartum, and worse systolic function (lower LVEF) from 6 months onward (LVMi: 77.61g/m^2^ [61.62-98.79] versus 65.11g/m^2^ [55.42-71.29], p=0.04; RWT: 0.48 [0.41-0.54] versus 0.45 [0.34-0.44], p=0.01; LVEF: 61.0% [60.0-62.0] versus 63.0% [60.5-64.0], p=0.03; Supplementary Table 6). In mixed-effects models accounting for repeated measures and adjusted for pre-existing hypertension, BP trajectories differed significantly across the postpartum period according to later hypertension status (Figure 4). Women who developed hypertension by 3-6 years demonstrated an attenuated decline in BP followed by a relative increase over time, following adjustment for pre-existing hypertension (systolic BP interaction: p=0.02; diastolic BP interaction: p=0.003; Figure 4), but no difference in change in systolic or diastolic function or pulse wave velocity over time. There was a trend towards early attenuated decline in LVMi in those who later developed hypertension (interaction, adjusted for pre-existing hypertension: p=0.05). There was no relationship between change in BMI over time and 3-6-year fibrosis or hypertension status, with or without adjustment for pre-existing hypertension.

**Figure 3:**
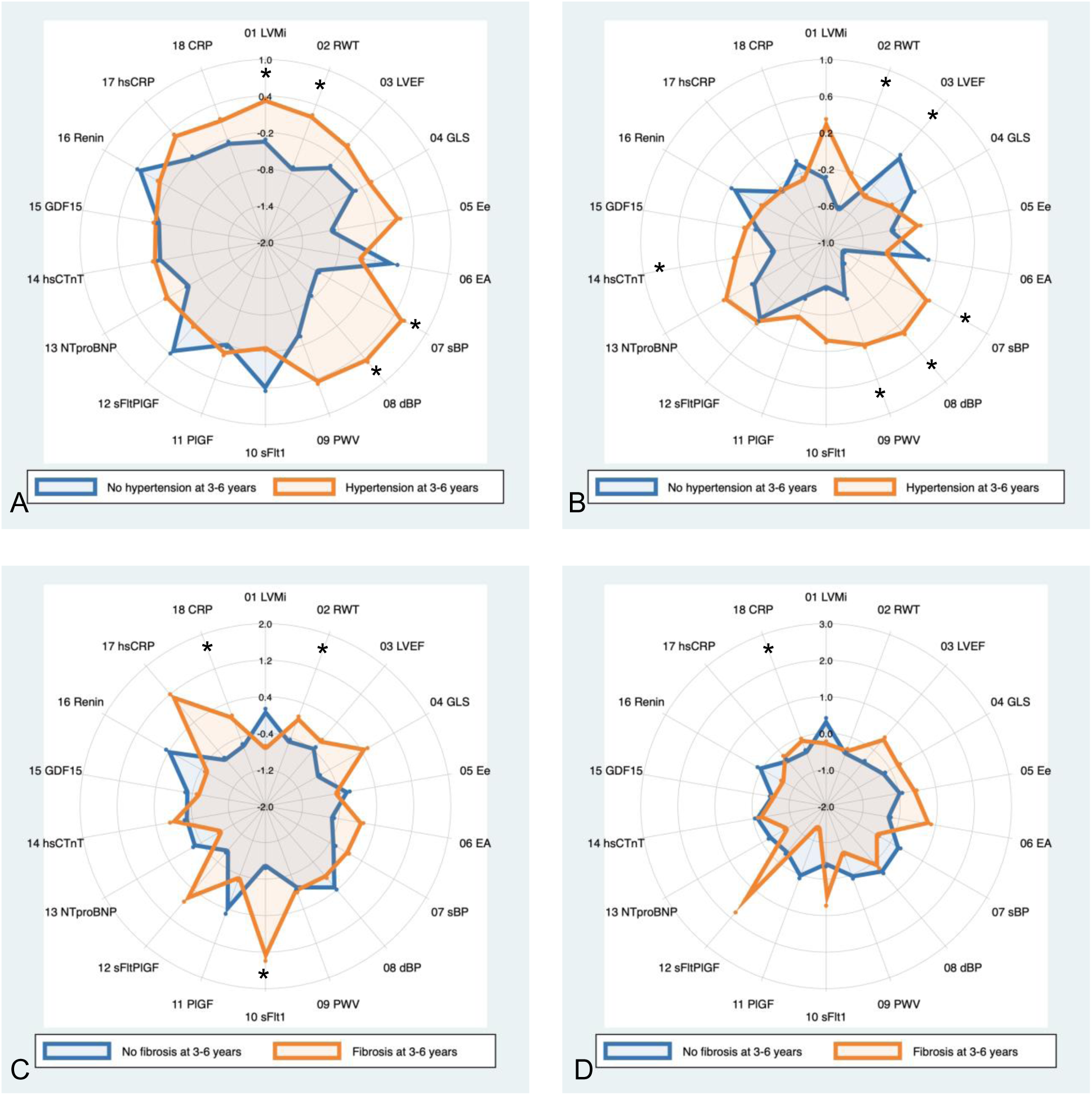
Differences in early postnatal phenotypes by cardiovascular morbidity at 3-6 years. Radar plots depict group-level mean standardized (z-score) values for blood pressure, cardiac structure and function, and circulating biomarkers measured in the early postnatal period. Values are scaled relative to the study population mean, with positive values indicating higher measurements and negative values indicating lower measurements. Shaded areas represent the reference distribution. Groups are stratified by cardiovascular outcomes at 3-6 years’ follow-up: (A) 6-week parameters by 3-6-year hypertension status; (B) 6-month parameters by 3-6-year hypertension status (C) 6-week parameters by 3-6-year myocardial fibrosis status; (D) 6-month parameters by 3-6-year myocardial fibrosis status. Abbreviations: BP, blood pressure; SBP, systolic blood pressure; DBP, diastolic blood pressure; LV, left ventricular; LVMI, left ventricular mass index; RWT, relative wall thickness; LVEF, left ventricular ejection fraction; GLS, global longitudinal strain; LA, left atrial; PWV, pulse wave velocity; NT-proBNP, N-terminal pro-B-type natriuretic peptide; hs-cTn, high-sensitivity cardiac troponin * Indicates statistical significance (p<0.05).

**Figure 4:**
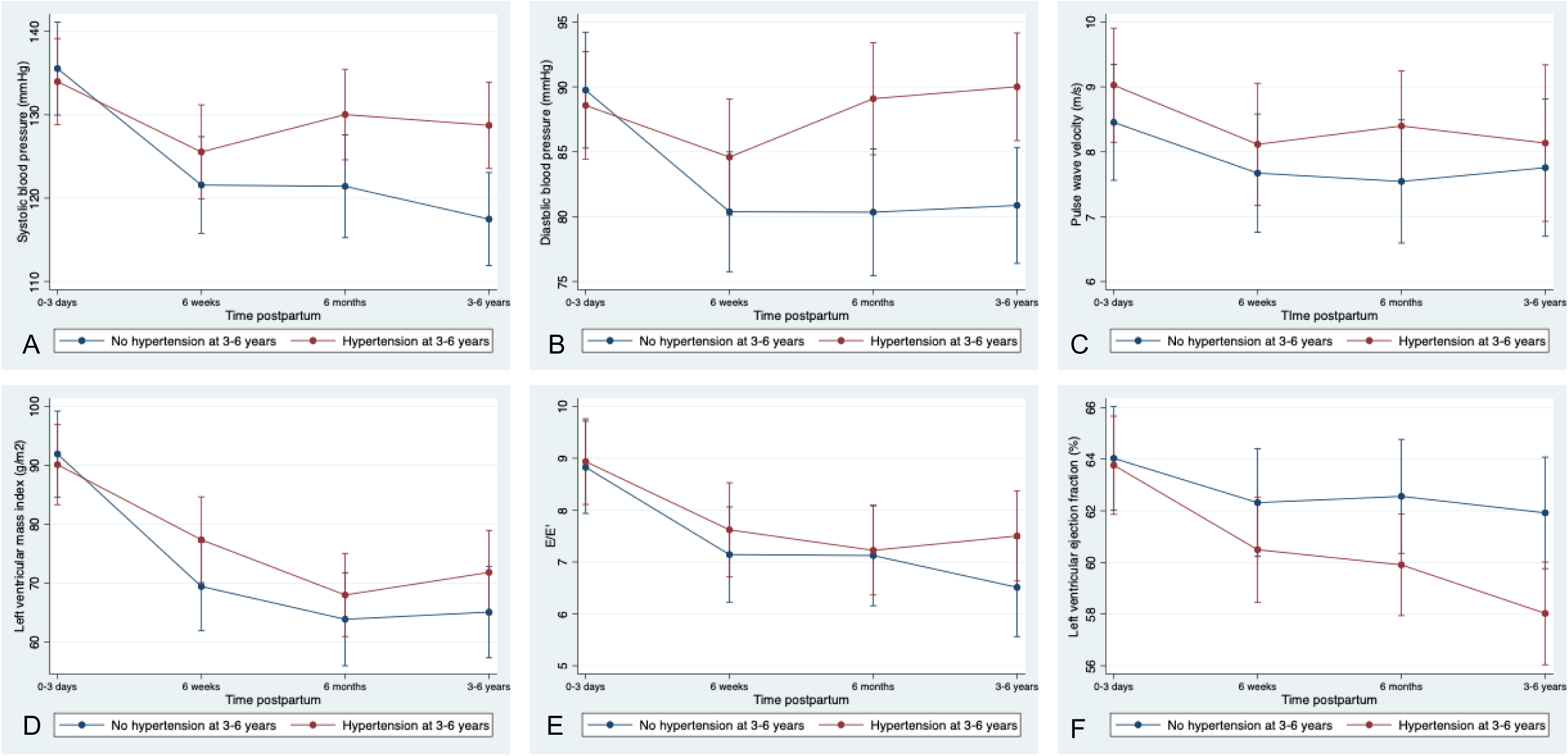
Change in cardiometabolic parameters over time by 3-6-year hypertension status. Change in cardiovascular parameters over time stratified by hypertension status at 3-6 years postpartum (hypertension n=24; no hypertension n=21). **(A)** systolic blood pressure, **(B)** diastolic blood pressure, **(C)** pulse wave velocity, **(D)** left ventricular mass index (LVMi), **(E)** diastolic function (E/E′), and **(F)** systolic function (left ventricular ejection fraction). Lines represent mean values and error bars represent standard errors. P values correspond to the time × hypertension status interaction term from mixed-effects regression models, with random effects to account for repeated measures within individuals and adjustment for pre-existing hypertension. LVMi, left ventricular mass index.

Among women who developed de novo hypertension by 3-6 years (10/31, 30%), BP was higher from 6 months (6-month systolic BP: 139.3 mmHg [127.3-144.7] versus 116.8 mmHg [112.8-120.3], p=0.03; 6-month diastolic BP: 95.3 mmHg [93.7-97.3] versus 76.2 mmHg [71.8-81.5], p=0.01; Supplementary Table 7).

Women who developed myocardial fibrosis by 3-6 years postpartum (7/20) had higher circulating sFlt and CRP concentrations from 6 weeks postpartum compared with women without fibrosis (6-week sFlt: 96.50 pg/mL [91.84-109.30] versus 84.20 pg/mL [77.92-91.92], p=0.03; 6-week CRP:10.00 mg/L [8.00-16.00] versus 1.00 [1.00-3.00], p=0.01; Supplementary Table 8). Regression analyses also demonstrated a positive linear association between serum sFlt levels and native T1 (a marker of myocardial fibrosis) at 3-6 years postpartum (r^2^= 0.43; p=0.002; Supplementary Figure 3) but not serum CRP levels and native T1 (log-transformed CRP: r^2^=0.07; p=0.29; Supplementary Figure 3).

## Discussion

In this longitudinal cohort of women with a history of preterm pre-eclampsia, we demonstrate a high prevalence of cardiovascular morbidity 3-6 years postpartum, including de novo hypertension and myocardial fibrosis. Overall, hypertension prevalence was more than fivefold higher than that reported in the UK population of reproductively aged women (53% versus 8%).^27^

Distinct adverse cardiovascular phenotypes were evident in the early postnatal period among those who later developed hypertension. Although attenuation by exclusion of women with pre-existing hypertension suggests contribution of pre-pregnancy status, persistence of differential postnatal BP trajectories, independent of pre-existing hypertension, indicates that postnatal cardiovascular recovery differences also contribute to subsequent risk.

Early postnatal sFlt was higher in those who later developed myocardial fibrosis. sFlt is released by the placenta and is a central mediator of endothelial dysfunction in pre-eclampsia. Persistently elevated sFlt levels, produced by endothelial cells and circulating monocytes,^28,29^ have been reported years after pre-eclampsia,^22,23^ and are associated with heart failure, renal disease, and adverse cardiovascular function.^22,30–32^ Experimental data show sFlt causes myocardial capillary rarefaction, impaired perfusion, and fibrosis.^33^ The association between sFlt and native T1 supports a mechanistic link via persistent angiogenic imbalance leading to microvascular dysfunction, myocardial hypoxia, and progressive interstitial fibrosis, rather than epicardial coronary atherosclerosis. Although underpowered to establish a definitive relationship between pre-eclampsia severity and later myocardial fibrosis, in this cohort, women with early-onset (<34 weeks) disease had a higher risk of fibrosis, suggesting a potential dose-effect, in line with Joubert et al.’s findings.^34^ In our study, variability in timing of the 0-3-day visit and absence of antenatal angiogenic marker measurements precluded assessment of peak pregnancy sFlt levels or trajectories from pregnancy into the postnatal period and therefore limited the ability to directly link antenatal angiogenic burden with later myocardial fibrosis.

Higher postnatal CRP was also associated with later fibrosis, supporting a role for sustained low-grade inflammation in adverse cardiac remodelling. Pre-eclampsia is characterized by systemic inflammation,^35–37^ and elevated CRP has been shown to persist decades postpartum, and to be associated with increased cardiovascular risk.^38,39^ Experimental data suggest CRP may exert pro-fibrotic effects through activation of angiotensin II and downstream TFG-ß/SMAD signalling pathways, further supporting inflammation as an active contributor to fibrotic remodelling rather than a passive marker.^40,41^ This is in line with Rodrigues et al.’s findings of BP-independent mechanisms driving myocardial fibrosis in hypertensive heart disease.^42^

Obesity is also characterized by chronic low-grade inflammation, with elevated CRP and excess adipose tissue that stores and secretes cytokines, including TNF-a and IL-6, both of which have been implicated in adverse cardiac remodelling and fibrosis.^43^ This provides a plausible biological explanation for the observed trend towards an association between higher booking BMI and subsequent myocardial fibrosis in this cohort.

Early postnatal BP and BMI identified women at increased risk of future hypertension 3-6 years before clinical diagnosis. Kitt et al. have demonstrated that proactive BP management in the early postnatal period can result in sustained improvement in BP control.^44,45^ Weight loss, including via glucagon-like peptide-1 [GLP-1] agonists, offers additional prevention potential but requires further safety data in the postnatal setting.^46,47^

Together these findings support a model in which early postnatal cardiovascular phenotype and biomarker profiles following pre-eclampsia may identify women at greatest risk of later cardiovascular morbidity.

Key strengths of this study include its prospective, longitudinal design, detailed cardiovascular phenotyping during pregnancy and the early postnatal period, and the use of multimodal cardiovascular assessment, including CMR. The cohort was well characterized clinically, enabling exploration of early postnatal trajectories in relation to later cardiovascular morbidity. Several limitations should be acknowledged. The absence of a normotensive comparator group limits direct attribution of observed abnormalities to pre-eclampsia rather than background population risk. The modest sample size also limited statistical power, particularly for subgroup analyses. In particular, the small number of women exposed to postnatal enalapril precluded meaningful assessment of the long-term impact of early antihypertensive treatment with subsequent cardiovascular outcomes.

In addition, pre-pregnancy cardiovascular phenotyping was not available, and therefore it is not possible to determine whether the cardiovascular abnormalities observed in the early postnatal period were present prior to pregnancy or emerged as a consequence of pre-eclampsia. As such, while the findings support early postnatal divergence in cardiovascular phenotype, causality and temporal directionality cannot be definitively established.

Finally, there was some mismatch in timing between bedside cardiovascular assessments (including BP measurement, blood sampling and echocardiography) and CMR imaging at 3-6 years postpartum, which limited direct cross-modal correlation of contemporaneous measures. Notably, despite this interval, a strong linear association between early postnatal circulating sFlt and later native T1 mapping was observed, underscoring the robustness and potential biological relevance of this finding.

## Perspectives

This study highlights preterm pre-eclampsia as a pivotal cardiovascular event, with adverse phenotypes emerging in the early postnatal period and persisting years later. The mechanistic association between postnatal sFlt and later myocardial fibrosis suggests that sustained endothelial dysfunction following preterm pre-eclampsia, rather than pre-pregnancy susceptibility alone, drives long-term risk. These findings support a shift towards earlier and more structured postnatal cardiovascular surveillance and intervention in this high-risk population.

Several research priorities emerge. Larger longitudinal studies are needed to characterize angiogenic and inflammatory trajectories in detail, and determine whether early postnatal interventions modify long-term cardiovascular risk. Adequately powered randomized trials should evaluate whether early antihypertensive therapy influences subsequent cardiovascular remodelling or clinical outcomes. Mechanistic studies examining sustained sFlt exposure effects on endothelial and myocardial cells may clarify causal pathways and identify novel therapeutic targets for cardiovascular disease prevention after pre-eclampsia.

## Conclusion

The findings of this study indicate that cardiometabolic morbidity is prevalent following preterm pre-eclampsia with adverse cardiovascular phenotypes evident in the early postnatal period. A sustained inflammatory state and anti-angiogenic imbalance may contribute to the development of myocardial fibrosis. Elevated BP in the early postnatal period was associated with hypertension at 3-6 years, suggesting that tight BP control may have an important role in postnatal care. Further clarification through larger observational and interventional studies is required to determine whether targeting angiogenic imbalance, inflammation, and early postnatal BP control can modify long-term cardiovascular risk following preterm pre-eclampsia.

## Novelty and relevance

Our findings extend the concept of pre-eclampsia as an early cardiovascular stress test, by suggesting that subsequent risk may also reflect differences in postnatal cardiovascular recovery, rather than baseline susceptibility alone.

### What is new?

- There is a high prevalence of de novo hypertension and myocardial fibrosis 3-6 years following preterm pre-eclampsia
- Cardiovascular risk after preterm pre-eclampsia may reflect impaired postnatal recovery, rather than baseline susceptibility alone
- Postnatal sFlt and inflammatory markers are associated with later myocardial fibrosis

### What is relevant?

- Hypertension prevalence after preterm pre-eclampsia is more than fivefold higher than in the general population
- Early postnatal blood pressure trajectories are altered in those who later develop chronic hypertension
- Study findings support mechanisms linking pre-eclampsia and cardiovascular disease, including postnatal hypertension, endothelial dysfunction and inflammation

### Clinical/pathophysiological implications?

- Postnatal BP and BMI assessment may enable timely identification of women at high risk of future hypertension
- Persistent angiogenic imbalance and inflammation may contribute to microvascular dysfunction and fibrotic myocardial remodelling
- Targeted postnatal surveillance and intervention represent potential strategies to reduce long-term cardiovascular risk after preterm pre-eclampsia

## Data Availability

The data underlying this study are not publicly available due to participant confidentiality and data protection regulations. De-identified data may be made available from the corresponding author upon reasonable request, subject to appropriate approvals and data sharing agreements.

## Acknowledgements

This work was supported by funding from the Medical Research Council (MRC), Wellbeing of Women and The Dowager Countess Eleanor Peel Trust, and Tommy’s Charity. LO is funded by NIHR as a Clinical Lecturer. CAM, Advanced Fellowship, NIHR301338 is funded by the National Institute for Health and Care Research (NIHR). The views expressed in this publication are those of the authors and not necessarily those of the NIHR, NHS or the UK Department of Health and Social Care. CAM acknowledges support from the University of Manchester British Heart Foundation Research Excellence Award (RE/24/130017) and the NIHR Manchester Biomedical Research Centre (NIHR203308). CAM has participated on advisory boards/consulted for AstraZeneca, Boehringer Ingelheim and Lilly Alliance, Novartis and PureTech Health, serves as an advisor for HAYA Therapeutics, has received speaker fees from AstraZeneca, Boehringer Ingelheim and Novo Nordisk, conference attendance support from AstraZeneca, and research support from Amicus Therapeutics, AstraZeneca, Guerbet Laboratories Limited, Roche and Univar Solutions B.V.

## References

1. Shih K, Herz N, Sheikh A, O’Neill C, Carter P, Anderson M. Economic burden of cardiovascular disease in the United Kingdom. Eur Hear J - Qual Care Clin Outcomes [Internet]. 2025 Aug 11 [cited 2026 Jan 22];11(5):678–90. Available from: 10.1093/ehjqcco/qcaf011

2. Vervoort D, Wang R, Li G, Filbey L, Maduka O, Brewer LPC, et al. Addressing the Global Burden of Cardiovascular Disease in Women: JACC State-of-the-Art Review. J Am Coll Cardiol [Internet]. 2024 Jun 25 [cited 2026 Jan 22];83(25):2690–707. Available from: /doi/pdf/10.1016/j.jacc.2024.04.028?download=true

3. Tayal U, Pompei G, Wilkinson I, Adamson D, Sinha A, Hildick-Smith D, et al. Advancing the access to cardiovascular diagnosis and treatment among women with cardiovascular disease: a joint British Cardiovascular Societies’ consensus document. Heart [Internet]. 2024 Nov 1 [cited 2026 Jan 22];110(22):e3–15. Available from: https://heart.bmj.com/content/110/22/e3

4. Wu R, Wang T, Gu R, Xing D, Ye C, Chen Y, et al. Hypertensive Disorders of Pregnancy and Risk of Cardiovascular Disease-Related Morbidity and Mortality: A Systematic Review and Meta-Analysis. Cardiology [Internet]. 2020 Oct 1 [cited 2020 Nov 25];145(10):633–47. Available from: https://www.karger.com/Article/FullText/508036

5. Leon LJ, McCarthy FP, Direk K, Gonzalez-Izquierdo A, Prieto-Merino D, Casas JP, et al. Preeclampsia and Cardiovascular Disease in a Large UK Pregnancy Cohort of Linked Electronic Health Records: A CALIBER Study. Circulation. 2019;140(13):1050–60.

6. Inversetti A, Pivato CA, Cristodoro M, Latini AC, Condorelli G, Di Simone N, et al. Update on long-term cardiovascular risk after pre-eclampsia: a systematic review and meta-analysis. Eur Hear J - Qual Care Clin Outcomes [Internet]. 2024 Jan 1 [cited 2025 May 5];10(1):4–13. Available from: https://pubmed.ncbi.nlm.nih.gov/37974053/

7. Arnott C, Nelson M, Alfaro Ramirez M, Hyett J, Gale M, Henry A, et al. Maternal cardiovascular risk after hypertensive disorder of pregnancy. Heart [Internet]. 2020 Dec 1 [cited 2020 Jul 21];106(24):1927–33. Available from: 10.1136/heartjnl-2020-316541

8. Smith GCS, Pell JP, Walsh D. Pregnancy complications and maternal risk of ischaemic heart disease: A retrospective cohort study of 129 290 births. Lancet [Internet]. 2001 Jun 23 [cited 2020 Jul 21];357(9273):2002–6. Available from: https://pubmed.ncbi.nlm.nih.gov/11438131/

9. Lykke JA, Langhoff-Roos J, Sibai BM, Funai EF, Triche EW, Paidas MJ. Hypertensive pregnancy disorders and subsequent cardiovascular morbidity and type 2 diabetes mellitus in the mother. Hypertension [Internet]. 2009 Jun [cited 2020 Jul 21];53(6):944–51. Available from: https://pubmed.ncbi.nlm.nih.gov/19433776/

10. Wikström AK, Haglund B, Olovsson M, Lindeberg SN. The risk of maternal ischaemic heart disease after gestational hypertensive disease. BJOG An Int J Obstet Gynaecol [Internet]. 2005 Nov [cited 2020 Jul 21];112(11):1486–91. Available from: https://pubmed.ncbi.nlm.nih.gov/16225567/

11. Magee LA, Brown MA, Hall DR, Gupte S, Hennessy A, Karumanchi SA, et al. The 2021 International Society for the Study of Hypertension in Pregnancy classification, diagnosis & management recommendations for international practice. Pregnancy Hypertens. 2022 Mar 1;27:148–69.

12. Inversetti A, Pivato CA, Cristodoro M, Latini AC, Condorelli G, Di Simone N, et al. Update on long-term cardiovascular risk after pre-eclampsia: a systematic review and meta-analysis. Eur Hear J - Qual Care Clin Outcomes [Internet]. 2024 Jan 12 [cited 2024 Apr 27];10(1):4–13. Available from: 10.1093/ehjqcco/qcad065

13. Wu P, Haththotuwa R, Kwok CS, Babu A, Kotronias RA, Rushton C, et al. Preeclampsia and Future Cardiovascular Health: A Systematic Review and Meta-Analysis. Circ Cardiovasc Qual Outcomes. 2017;10(2):e003497.

14. McCarthy FP, O’Driscoll J, Seed P, Brockbank A, Cox A, Gill C, et al. Planned delivery to improve postpartum cardiac function in women with preterm pre-eclampsia: the PHOEBE mechanisms of action study within the PHOENIX RCT. Effic Mech Eval [Internet]. 2021 Sep [cited 2022 Feb 15];8(12):1–28. Available from: https://www.ncbi.nlm.nih.gov/books/NBK574014/

15. Craici I, Wagner S, Garovic VD. Review: Preeclampsia and future cardiovascular risk: formal risk factor or failed stress test? Ther Adv Cardiovasc Dis [Internet]. 2008 [cited 2026 Feb 5];2(4):249–59. Available from: /doi/pdf/10.1177/1753944708094227?download=true

16. Thakkar A, Hailu T, Blumenthal RS, Martin SS, Harrington CM, Yeh DDF, et al. Cardio-Obstetrics: the Next Frontier in Cardiovascular Disease Prevention. Curr Atheroscler Rep [Internet]. 2022 Jul 1 [cited 2026 Feb 5];24(7):493–507. Available from: https://link.springer.com/article/10.1007/s11883-022-01026-6

17. Thilaganathan B, Kalafat E. Cardiovascular System in Preeclampsia and Beyond. Hypertens (Dallas, Tex 1979) [Internet]. 2019 Mar 1 [cited 2026 Feb 5];73(3):522. Available from: https://pmc.ncbi.nlm.nih.gov/articles/PMC6380450/

18. Bellamy L, Casas JP, Hingorani AD, Williams DJ. Pre-eclampsia and risk of cardiovascular disease and cancer in later life: Systematic review and meta-analysis. Br Med J [Internet]. 2007 Nov 10 [cited 2020 Jul 21];335(7627):974–7. Available from: http://www.bmj.com/cgi/doi/10.1136/bmj.39335.385301.BE

19. Brown MC, Best KE, Pearce MS, Waugh J, Robson SC, Bell R. Cardiovascular disease risk in women with pre-eclampsia: Systematic review and meta-analysis. Eur J Epidemiol. 2013;28(1):1–19.

20. Heida KY, Franx A, Van Rijn BB, Eijkemans MJC, Boer JMA, Verschuren MWM, et al. Earlier Age of Onset of Chronic Hypertension and Type 2 Diabetes Mellitus After a Hypertensive Disorder of Pregnancy or Gestational Diabetes Mellitus. Hypertens (Dallas, Tex 1979) [Internet]. 2015 Dec 1 [cited 2026 Mar 7];66(6):1116–22. Available from: https://pubmed.ncbi.nlm.nih.gov/26459420/

21. Countouris ME, Bello NA. Advances in Our Understanding of Cardiovascular Diseases After Preeclampsia. Circ Res [Internet]. 2025 Mar 14 [cited 2026 Mar 7];136(6):583–93. Available from: /doi/pdf/10.1161/CIRCRESAHA.124.325581?download=true

22. Wolf M, Hubel CA, Lam C, Sampson M, Ecker JL, Ness RB, et al. Preeclampsia and future cardiovascular disease: Potential role of altered angiogenesis and insulin resistance. J Clin Endocrinol Metab [Internet]. 2004 Dec [cited 2020 Jul 29];89(12):6239–43. Available from: https://pubmed.ncbi.nlm.nih.gov/15579783/

23. Kvehaugen AS, Dechend R, Ramstad HB, Troisi R, Fugelseth D, Staff AC. Endothelial function and circulating biomarkers are disturbed in women and children after preeclampsia. Hypertension [Internet]. 2011 Jul [cited 2020 Jul 30];58(1):63–9. Available from: https://pubmed.ncbi.nlm.nih.gov/21606387/

24. Shahul S, Medvedofsky D, Wenger JB, Nizamuddin J, Brown SM, Bajracharya S, et al. Circulating antiangiogenic factors and myocardial dysfunction in hypertensive disorders of pregnancy. Hypertension [Internet]. 2016 Jun 1 [cited 2020 Jul 29];67(6):1273–80. Available from: https://pubmed.ncbi.nlm.nih.gov/27113052/

25. Ormesher L, Higson S, Luckie M, Roberts SA, Glossop H, Trafford A, et al. Postnatal Enalapril to Improve Cardiovascular Function Following Preterm Preeclampsia (PICk-UP):: A Randomized Double-Blind Placebo-Controlled Feasibility Trial. Hypertension [Internet]. 2020 Oct 5 [cited 2020 Oct 6];76(6):1828–37. Available from: http://www.ncbi.nlm.nih.gov/pubmed/33012200

26. Ormesher L, Higson S, Luckie M, Roberts SA, Glossop H, Trafford A, et al. Postnatal cardiovascular morbidity following preterm pre-eclampsia: An observational study. Pregnancy Hypertens. 2022;(30):68–81.

27. NHS England. Adults’ health: Hypertension - NHS England Digital01 Jan 2021 to 31 Mar 2022 - [Internet]. 2023 [cited 2025 May 8]. Available from: https://digital.nhs.uk/data-and-information/publications/statistical/health-survey-for-england/2021-part-2/adult-health-hypertension

28. Di Marco GS, Reuter S, Hillebrand U, Amler S, König M, Larger E, et al. The soluble VEGF receptor sFlt1 contributes to endothelial dysfunction in CKD. J Am Soc Nephrol [Internet]. 2009 Oct [cited 2020 Oct 12];20(10):2235–45. Available from: /pmc/articles/PMC2754110/?report=abstract

29. Weissgerber TL, Rajakumar A, Myerski AC, Edmunds LR, Powers RW, Roberts JM, et al. Vascular pool of releasable soluble VEGF receptor-1 (sFLT1) in women with previous preeclampsia and uncomplicated pregnancy. J Clin Endocrinol Metab [Internet]. 2014 [cited 2026 Feb 6];99(3):978–87. Available from: https://pubmed.ncbi.nlm.nih.gov/24423299/

30. Gruson D, Hermans MP, Ferracin B, Ahn SA, Rousseau MF. Sflt-1 in heart failure: relation with disease severity and biomarkers. Scand J Clin Lab Invest [Internet]. 2016 Jul 3 [cited 2020 Aug 3];76(5):411–6. Available from: https://www.tandfonline.com/doi/abs/10.1080/00365513.2016.1190863

31. Hammadah M, Georgiopoulou V V., Kalogeropoulos AP, Weber M, Wang X, Samara MA, et al. Elevated Soluble Fms-Like Tyrosine Kinase-1 and Placental-Like Growth Factor Levels Are Associated with Development and Mortality Risk in Heart Failure. Circ Hear Fail [Internet]. 2016 Jan 1 [cited 2020 Aug 3];9(1):e002115. Available from: /pmc/articles/PMC4692173/?report=abstract

32. Draker N, Torry DS, Torry RJ. Placenta growth factor and sFlt-1 as biomarkers in ischemic heart disease and heart failure: a review. Biomark Med [Internet]. 2019 [cited 2026 Feb 6];13(9):785–99. Available from: https://pubmed.ncbi.nlm.nih.gov/31157982/

33. Di Marco GS, Kentrup D, Reuter S, Mayer AB, Golle L, Tiemann K, et al. Soluble Flt-1 links microvascular disease with heart failure in CKD. Basic Res Cardiol [Internet]. 2015 Apr 13 [cited 2020 Jun 23];110(3):30. Available from: https://pubmed.ncbi.nlm.nih.gov/25893874/

34. Joubert LH, Doubell AF, Langenegger EJ, Herrey AS, Bergman L, Bergman K, et al. Cardiac magnetic resonance imaging in preeclampsia complicated by pulmonary edema shows myocardial edema with normal left ventricular systolic function. Am J Obstet Gynecol [Internet]. 2022 Aug 1 [cited 2026 Jan 23];227(2):292.e1–292.e11. Available from: https://www.sciencedirect.com/science/article/abs/pii/S0002937822001806

35. Singh SB, Mahajan S, PS K, Mangla D, Singh SB, Mahajan S, et al. Study of C-reactive Protein Levels in Hypertensive Disorders of Pregnancy. Cureus [Internet]. 2024 Aug 15 [cited 2026 Jan 22];16(8). Available from: https://cureus.com/articles/236165-study-of-c-reactive-protein-levels-in-hypertensive-disorders-of-pregnancy

36. Parchim NF, Wang W, Iriyama T, Ashimi OA, Siddiqui AH, Blackwell S, et al. Neurokinin 3 receptor and phosphocholine transferase: Missing factors for pathogenesis of C-reactive protein in preeclampsia. Hypertension [Internet]. 2015 Feb 21 [cited 2026 Jan 22];65(2):430–9. Available from: /doi/pdf/10.1161/HYPERTENSIONAHA.114.04439?download=true

37. Rebelo F, Schlüssel MM, Vaz JS, Franco-Sena AB, Pinto TJP, Bastos FI, et al. C-reactive protein and later preeclampsia: Systematic review and meta-analysis taking into account the weight status. J Hypertens [Internet]. 2013 [cited 2026 Jan 22];31(1):16–26. Available from: https://journals.lww.com/jhypertension/fulltext/2013/01000/c_reactive_protein_and_later_preeclampsia_.3.aspx

38. Liu C, Li C. C-reactive protein and cardiovascular diseases: a synthesis of studies based on different designs. Eur J Prev Cardiol [Internet]. 2023 Oct 26 [cited 2026 Jan 22];30(15):1593–6. Available from: 10.1093/eurjpc/zwad116

39. Takvorian KS, Wang D, Courchesne P, Vasan RS, Benjamin EJ, Cheng S, et al. The Association of Protein Biomarkers With Incident Heart Failure With Preserved and Reduced Ejection Fraction. Circ Hear Fail [Internet]. 2023 Jan 1 [cited 2026 Jan 22];16(1):E009446. Available from: /doi/pdf/10.1161/CIRCHEARTFAILURE.121.009446?download=true

40. Leal CRV, Costa LB, Ferreira GC, Ferreira A de M, Reis FM, Simões e Silva AC. Renin-angiotensin system in normal pregnancy and in preeclampsia: A comprehensive review. Pregnancy Hypertens [Internet]. 2022 Jun 1 [cited 2026 Jan 22];28:15–20. Available from: https://pubmed.ncbi.nlm.nih.gov/35149272/

41. Zhang R, Zhang YY, Huang XR, Wu Y, Chung ACK, Wu EX, et al. C-reactive protein promotes cardiac fibrosis and inflammation in angiotensin II-induced hypertensive cardiac disease. Hypertension [Internet]. 2010 Apr 1 [cited 2026 Jan 22];55(4):953–60. Available from: /doi/pdf/10.1161/HYPERTENSIONAHA.109.140608?download=true

42. Rodrigues JCL, Amadu AM, Dastidar AG, Szantho G V., Lyen SM, Godsave C, et al. Comprehensive characterisation of hypertensive heart disease left ventricular phenotypes. Heart [Internet]. 2016 Oct 15 [cited 2026 Feb 6];102(20):1671–9. Available from: https://heart.bmj.com/content/102/20/1671

43. Meléndez GC, McLarty JL, Levick SP, Du Y, Janicki JS, Brower GL. Interleukin 6 mediates myocardial fibrosis, concentric hypertrophy, and diastolic dysfunction in rats. Hypertension [Internet]. 2010 Aug 1 [cited 2026 Jan 22];56(2):225–31. Available from: /doi/pdf/10.1161/HYPERTENSIONAHA.109.148635?download=true

44. Kitt JA, Fox RL, Cairns AE, Mollison J, Burchert HH, Kenworthy Y, et al. Short-Term Postpartum Blood Pressure Self-Management and Long-Term Blood Pressure Control: A Randomized Controlled Trial. Hypertens (Dallas, Tex 1979) [Internet]. 2021 Aug 1 [cited 2022 Nov 21];78(2):469. Available from: /pmc/articles/PMC8260340/

45. Cairns AE, Tucker KL, Leeson P, Mackillop LH, Santos M, Velardo C, et al. Self-management of postnatal hypertension the SNAP-HT trial. Hypertension [Internet]. 2018 Aug [cited 2020 Nov 17];72(2):425–32. Available from: https://www.ahajournals.org/doi/10.1161/HYPERTENSIONAHA.118.10911

46. Budini B, Luo S, Tam M, Stead I, Lee A, Akrami A, et al. Trajectory of weight regain after cessation of GLP-1 receptor agonists: a systematic review and nonlinear meta-regression. eClinicalMedicine [Internet]. 2026 Mar [cited 2026 Mar 24];0(0):103796. Available from: https://www.thelancet.com/action/showFullText?pii=S258953702600043X

47. Parker CH, Slattery C, Brennan DJ, le Roux CW. Glucagon-like peptide 1 (GLP-1) receptor agonists’ use during pregnancy: Safety data from regulatory clinical trials. Diabetes, Obes Metab [Internet]. 2025 Aug 1 [cited 2026 Mar 24];27(8):4102–8. Available from: /doi/pdf/10.1111/dom.16437

